# Functional analysis of genome-wide dataset from 17000 individuals identifies multiple candidate malaria resistance genes enriched in malaria pathogenic pathways

**DOI:** 10.1101/2020.08.15.20175471

**Authors:** Delesa Damena, Francis Agamah, Peter O. Kimathi, Ntumba E. Kabongo, Hundaol Girma, Wonderful T. Choga, Lemu Golassa, Emile R. Chimusa

## Abstract

Recent genome-wide association studies (GWASs) of severe malaria have identified several association variants. However, much about the underlying biological functions are yet to be discovered. Here, we systematically predicted plausible candidate genes and pathways from functional analysis of severe malaria resistance GWAS summary statistics (N = 17,000) meta-analyzed across eleven populations in malaria endemic regions. We applied positional mapping, expression quantitative trait locus (eQTL), chromatin interaction mapping and gene-based association analyses to identify candidate severe malaria resistance genes. We performed network and pathway analyses to investigate their shared biological functions. We further applied rare variant analysis to raw GWAS datasets (N = 11,000) of three malaria endemic populations including Kenya, Malawi and Gambia and performed various population genetic structures of the identified genes in the three populations and global populations.

Our functional mapping analysis identified 57 genes located in the known malaria genomic loci while our gene-based GWAS analysis identified additional 125 genes across the genome. The identified genes were significantly enriched in malaria pathogenic pathways including multiple overlapping pathways in erythrocyte-related functions, blood coagulations, ion channels, adhesion molecules, membrane signaling elements and neuronal systems. Our population genetic analysis revealed that the minor allele frequencies (MAF) of the single nucleotide polymorphisms (SNPs) residing in the identified genes are generally higher in the three malaria endemic populations compared to global populations. Overall, our results suggest that severe malaria resistance trait is attributed to multiple genes; highlighting the possibility of harnessing new malaria therapeutics that can simultaneously target multiple malaria protective host molecular pathways.

## Introduction

Malaria is still one of the global health problems with approximately 228 million cases and 405, 000 deaths in 2018 (1). African countries disproportionately carry the global burden of malaria accounting for 93% and 94% of cases and deaths, respectively (1). *P. falciparum* malaria is still one of the leading causes of child mortality in endemic regions, particularly in sub-Saharan Africa. According to the World Health Organization (WHO), malaria killed about 285,000 under five children in 2016 (2). About 10–20% of children who recover from severe malaria develop neurological sequalae and sub-optimal neuronal development (3). Severe malaria (SM) is defined as demonstration of asexual forms of the malaria parasites in the blood of a patient with a potentially fatal manifestation or complication of malaria in whom other diagnosis have been excluded (4). The SM complications include rapid progression to severe malarial anaemia (SMA), hypoglycaemia, cerebral malaria(CM), acidosis and death (4).

The global malaria eradication program, accelerated by the WHO led Roll Back Malaria (RBM) partnership, is largely focusing on scaling up the coverage of the available intervention strategies such as the distribution of long-lasting insecticide treated nets (LLINS), indoor residual insecticide spraying, intermittent treatment for pregnant women in high transmission settings and rapid diagnosis and effective treatments using artemisinin-based combination therapies (ACTs) (5). This led to the significant decline of malaria burden in many parts of the endemic regions. Despite the successes gained in reducing the global burden of malaria, the progress towards global malaria elimination is challenged by wide arrays of problems including decreased funding, lack of political commitments in some countries, emergence of drug resistant parasites and insecticide resistant mosquitoes and lack of effective vaccine among others (6).

*P. falciparum* has a complex life cycle alternate between vertebrate and female *Anopheles mosquito*. During its blood meal, the infected mosquitoes inoculates the transmissive form the parasite, the sporozoites, in to human skin. From the skin the sporozoites enter in to the blood circulation or up-taken by the lymphatic system and invade liver (7). After maturation, the parasite buds off the hepatocytes and released in to the circulation in the form of merozomes containing hundreds of thousands of merozoites that infect erythrocytes (7). The erythrocyte stage also called blood stage lifecycle is complex multi-step process that involve repeated invasion, growth, replication and egress events (8). To create favourable environment for its survival and growth, the parasite remodels the infected erythrocyte (IE). The remodelling events include creation of a parasitophorous vacuolar membrane (PVM) that surrounds the parasite and modification of antigenic and structural properties of the IE by transporting a protein called *P. falciparum erythrocyte membrane protein 1 (PfEMP1)* to the surface (8). The clinical symptoms of the SM have been linked to the blood stage life cycle (8).

Even though SM is one of the commonest reasons for admission to hospital and is a major cause of hospital death in children aged 1–5 years in endemic areas, it constitutes only a small subset (1–2%) of the infected children as the majority of malaria infections is mild (9). It has been shown that such clinical variations is partly attributable to human genetic factors (10, 11). Thus, a comprehensive understanding of the human genetic causes of variation in malaria clinical outcome may potentially provide clues to design new intervention strategies such as therapeutics and vaccines which can facilitate the global malaria eradication program (12, 13).

Aiming at shedding more light to the genetic basis of severe *P. falciparum* malaria, several genome-wide association studies (GWASs) have been conducted in diverse malaria endemic populations over the last decade (14–18). The GWASs have replicated some of the well-known malaria susceptibility genomic risk loci including sickle cell (*HBB*) and *ABO* blood group loci and identified new variants in *ATP2B4* and Glycophorin regions (14–18). Due to the single-marker testing approach commonly used, single SNP-based GWASs may miss candidate variants with weak genetic effects and therefore, combining all effects from multiple variants within a gene and deconvoluting the interactions between genes underlying the malaria resistance trait may provide important insights in to underlying genetics (19). Today, a number of gene-based and pathway-level statistical analytic methods have recently been developed and successfully implemented in complex disease studies (20–23). These methods integrate functional information from advanced biological databases including the Genotype-Tissue Expression (GTEx)(24), Encyclopedia of DNA Elements (ENCODE) (25), Roadmap Epigenomics Project (26) and chromatin interaction information (27). Owing to the fact that direct functional follow-up of several candidate causal variants and genes is expensive, application of such computational method to prioritize genes and their respective biological pathways are proven to be useful in complex diseases studies (20). Furthermore, gene and gene-set analysis can improve the study power by aggregating the joint effects of weakly associated markers, a common challenge in a polygenic trait studies (28).

Here, we implement several gene-set, pathway and network analytic methods on summary statistics of severe malaria GWAS from 17,000 individuals meta-analysed across eleven populations and systematically predicted plausible genes and pathways. We further performed rare variant analysis on raw GWAS dataset (N = ∼11,000) of Kenya, Gambia and Malawi populations. Finally, we performed population genetic structure analysis of the identified genes in the three malaria endemic countries and across global populations. Established over the course of long coevolution time, blood stage life cycle of the parasite constitutes the most extensive interplay between host and parasite genomes which leads to the clinical symptoms of SM. Therefore, our results suggest that severe malaria resistance is polygenic and attributed to multiple genes aggregated in pathogenic pathways linked to the erythrocyte stage lifecycle of *P. falciparum*.

## Results

### Functional mapping and annotations

We applied three functional mapping strategies to the severe malaria GWAS summary statistics (N = 17,000 samples, ∼17 million SNPs) meta-analysed across 11 malaria endemic populations in Africa, Asia and Oceania (see **Material and Methods**). We identified 19 lead SNPs out of 69 significant SNPs across 6 genomic loci (**Supplementary Data 1–3**). The genomic locus was defined as the region that contain independent lead SNPs and nominally significant SNPs (p< 0.05) in linkage dis-equilibrium (LD) block with lead SNPs. An independent significant SNP was defined as a genome-wide significant SNP (P-value < 5e-8) within the genomic boundary of LD threshold of r^2^ > 0.6. Lead SNPs were selected from independent significant SNPs at LD threshold of r^2^ > 0.1. SNPs in close proximity (< 250 kb) were considered as a single locus and thus, each genomic locus can contain multiple independent significant SNPs and lead SNPs. These SNPs were significantly enriched in ncRNA-intronic, intronic, intergenic and ncRNA-exonic regions (**Fig1.A**).

**Fig.1.**
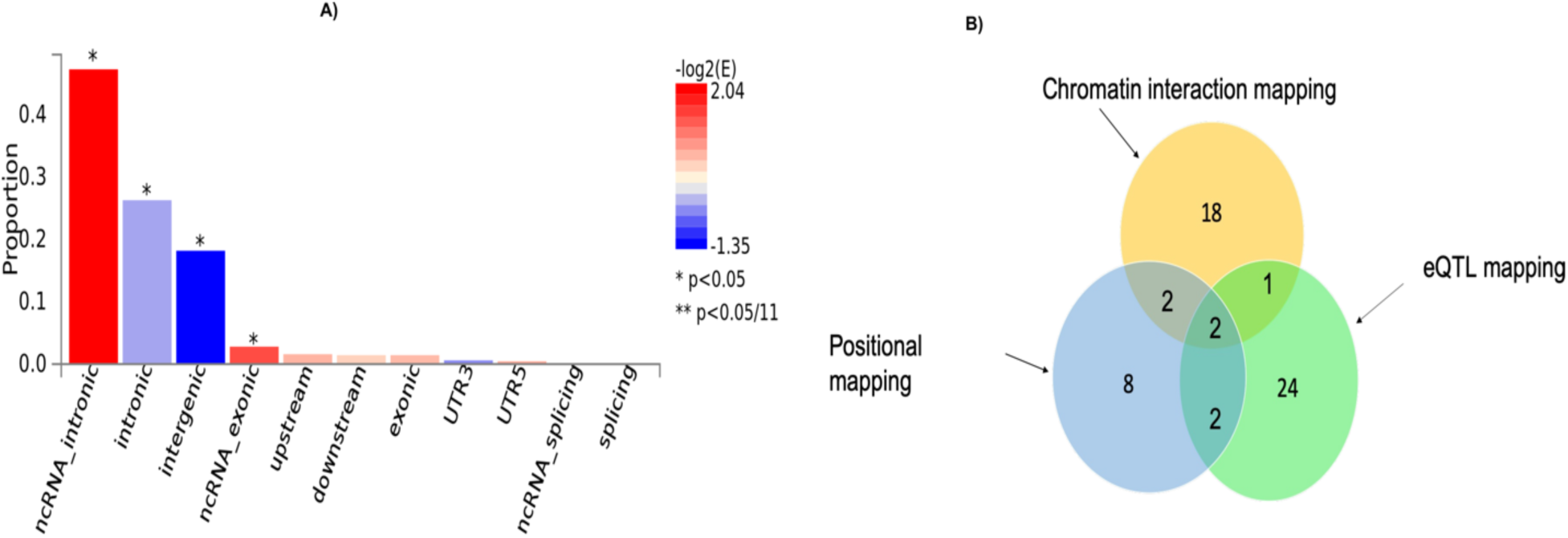
**A)** Proportion of malaria resistance GWAS SNPs in different genomic annotation categories. **B)** The number of genes identified by each of the three functional mapping strategies including positional mapping, eQTL and chromatin interactions. The intersection sections of the circles depict the number overlapped genes between the respective mapping strategies.

Our functional mapping strategies yielded a total of 57 protein-coding genes (**Table 1, Supplementary Data 4**). These include 29, 23 and 14 genes identified by eQTL mapping, chromatin interaction mapping and positional mapping, respectively (**Fig1.B**). Two genes including *ATP2B4* and *HBD* were identified by all the three gene mapping strategies while five genes including *GYPB, HBG2, TRIM6-TRIM34, OR51F2* and *TRIM68* were predicted by two of the three mapping strategies (**Supplementary Data 4**).

**Table 1.**
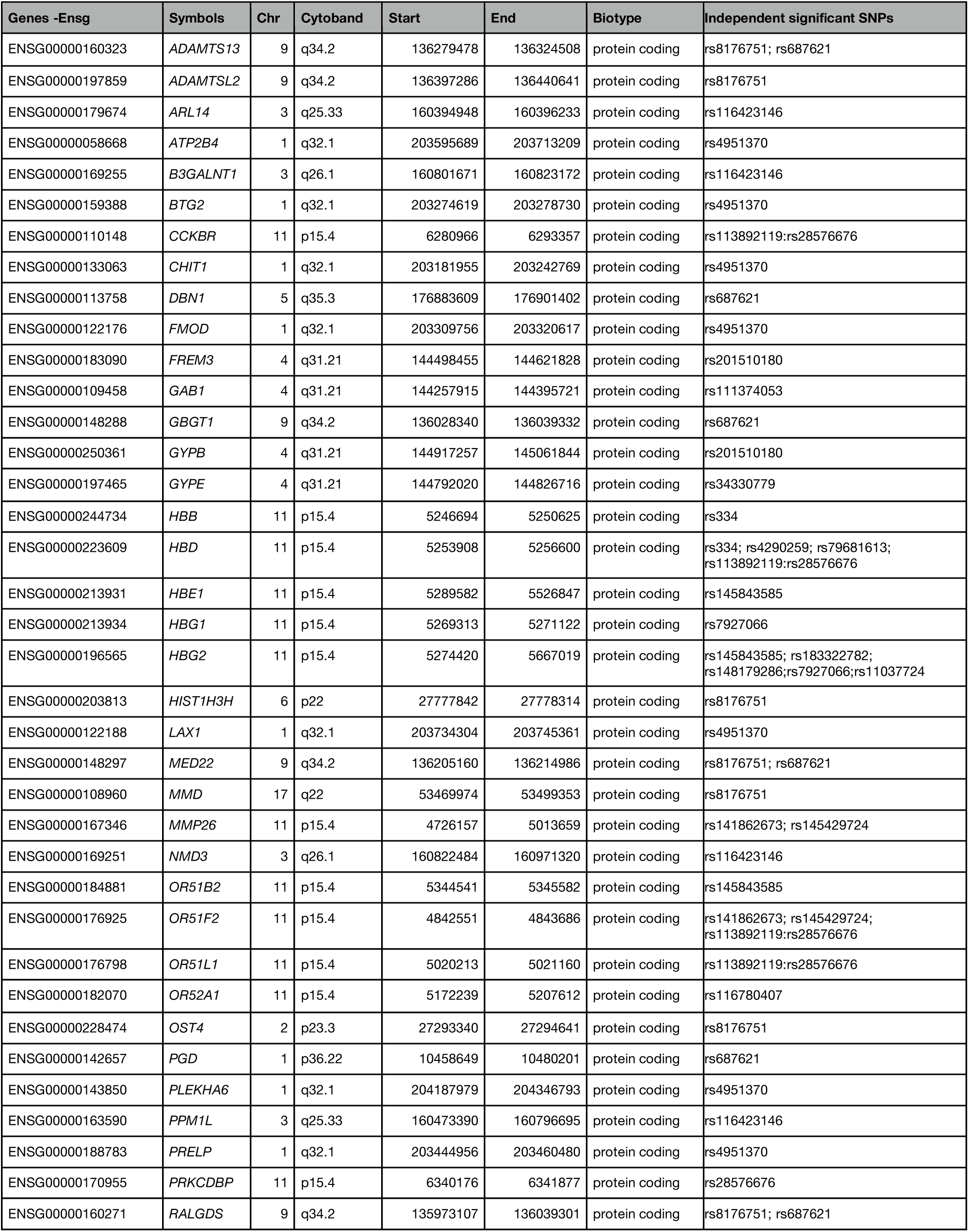
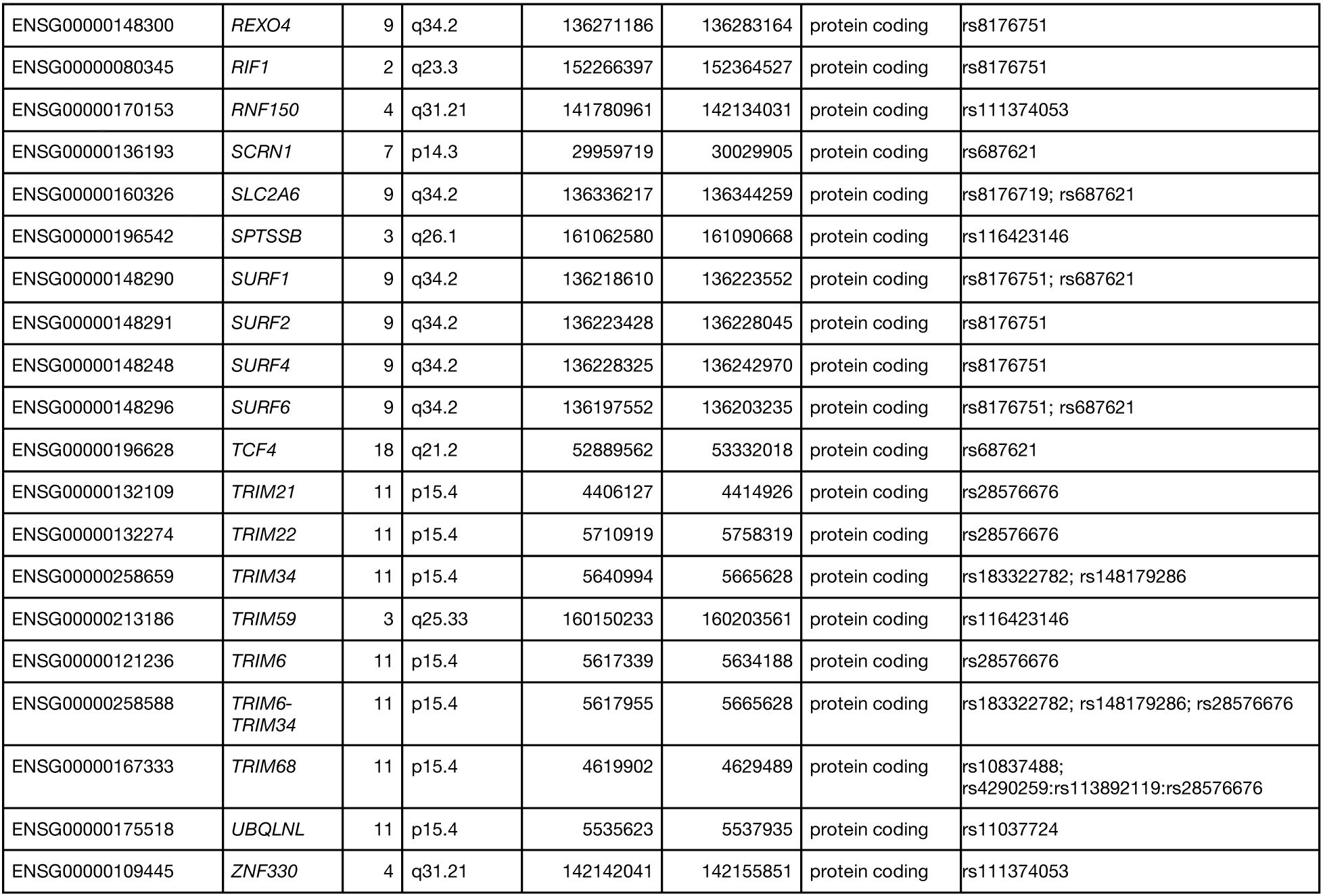
Fifty-seven severe malaria resistance candidate genes identified by eQTL mapping, chromatin interaction mapping and positional mapping strategies implemented in FUMA.

The identified genes were enriched in five cytogenic positions including 11p15 (p = 2.65e-18), chr9q34 (p = 4.63e-9), chr4p31(p = 4.6e-8), chr1q32 (p = 2.6e-7), chr3q26 (p = 7.97e-4) (S1 Table). We noted that the majority (33%) of the identified genes were clustered on 11p15 (**Fig 2**). These include beta globin gene cluster: *HBB, HBD, HBG1, HBG2* and *HBE1;* Tripartite motif-containing (*TRIM)* family genes including *TRIM68, TRIM21*; and genes involved in olfactory receptors and G protein-coupled signalling (GPCR) such as: *CCKR OR51F2* and *OR51L* (**Supplementary Table 1**). About two third (13/19) of the genes in this locus are in eQTL and chromatin interactions (**Supplementary Data 4 and Supplementary Fig 1)**.

All the implicated genes in chr9q34 locus are located outside the genomic risk locus and were identified by eQTL mapping (**Supplementary Data 4, Supplementary Fig 2)**. These include surfeit gene cluster such as: *SURF2, SURF4, MED22* and *SURF6;* a metalloprotease gene*, ADAMTS13;* and a gene encoding *ORS* blood group system coding gene (*GBGT1)*. In the remaining enriched cytogenic positions, the known genes including *ATP2B4* (chr1q32); *FREM3, GYPE* and *GYPB* (chr4p31) were replicated. Other notable genes include *BTG2* a tumour suppressor gene on chr1q32 and *B3GALNT1* on chr3q26 (**Supplementary Data 4**).

**Fig. 2.**
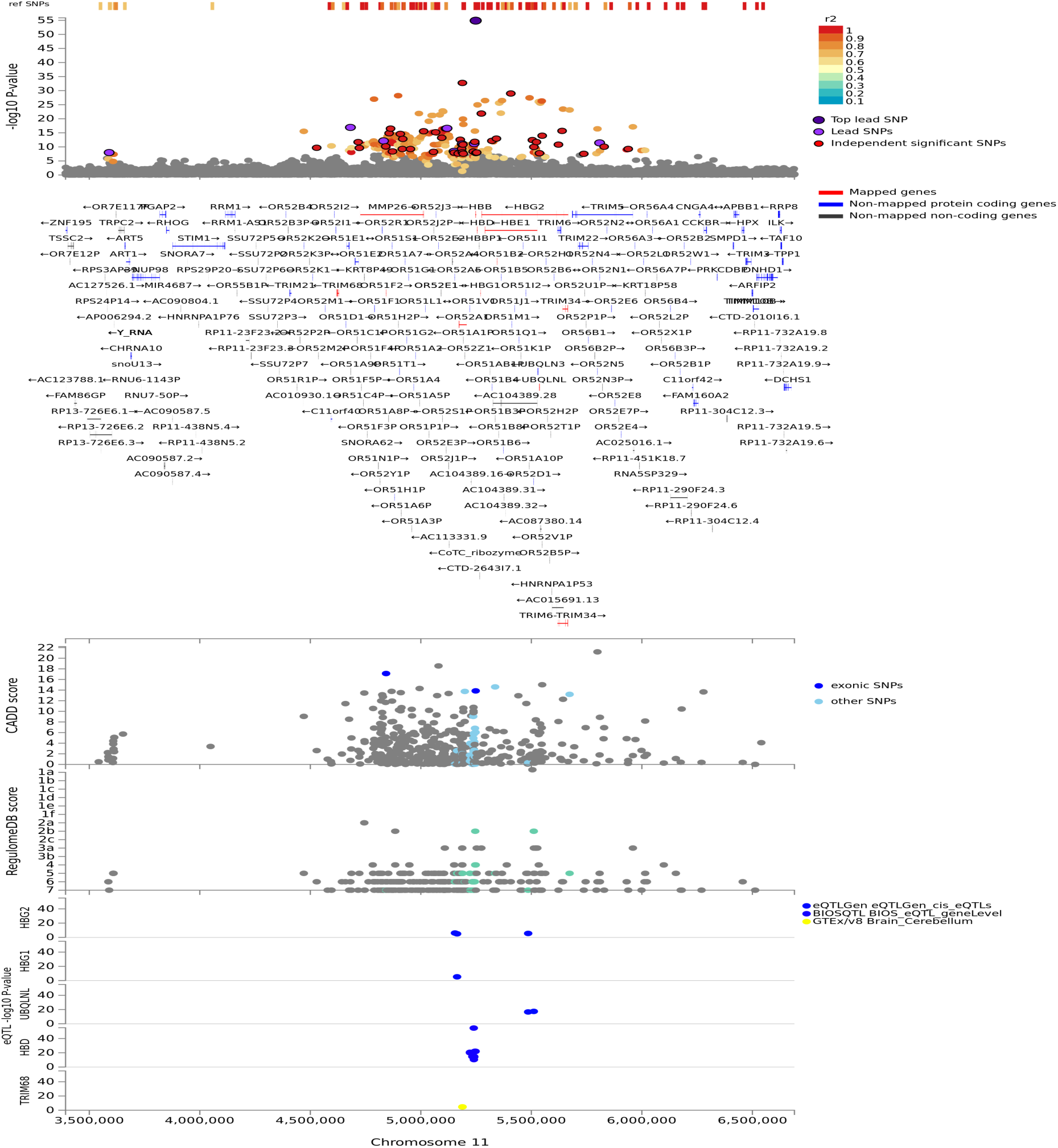
Regional plot of severe malaria susceptibility GWAS locus on chromosome 11. Non-GWAS-tagged SNPs are shown at the top of the plot as rectangles since they do not have a P-value from the GWAS. Prioritized genes are highlighted in red. eQTLs are plotted per gene and coloured based on tissue types. CADD score, RegulomeDB score and eQTLs, SNPs which are not mapped to any gene are coloured grey.

### Candidate genes identified by gene-based GWAS analysis

Taking the polygenic nature of severe malaria resistance trait in to consideration (11), we applied a pathway scoring algorithm(Pascal) (22) method that aggregates all SNPs within a gene and capture polygenic effects at the gene level (**see Material and Methods**). The Pascal analysis replicated 13 genes that were identified by our functional annotation methods in malaria genomic risk loci (**Supplementary Table 2**) and identified 125 additional genes across the genome (**Supplementary Data 5**). The genes with top scores outside genomic risk loci include *CSMD1* (p = 1.58e-12) on chr8p23.2 and *RBFOX1* (p = 9.76e-11) on chr16p13.3. *CSMD1* is an important regulator of complement activation and inflammation (29, 30) while *RBFOX1* encodes for an mRNA-splicing factor linked to autism spectrum disorders (31). A previous study in Tanzanian population reported association of variants in *RBFOX* gene with SM (17).

Other important genes identified by gene-based GWASs include neural adhesion molecules including *CNTN4* (p = 3.88e-9) on chr3p26.3-p26.2; *PCSK5*(p = 2.88e-11) on chr9q21.13; *CDH13* (p = 4.19e-8) on chr16q23.3) and *TMEM132* (p = 2.18e-8) on chr17q12 (**Supplementary Data 5**). These genes were reported to be linked to autism spectrum disorders and other neurodevelopmental conditions (32). Furthermore, protein kinases including *FLT4* (p = 9.96e-8) on 5q35.3; and *PTPRT* (p = 4.92e-7) on chr20q12-q13 and *PRKG1* ((p = 1.2e-6) on 10q11.2-q21.1 were among the genes with top scores (**Supplementary Data 5**). *PTPRT* is a tyrosine phosphatase receptor involved in STAT3 pathway and was recently reported to be associated with mild malaria susceptibility in Benin populations (33). *PRKG1* is a cyclic guanosine monophosphate (GMP) dependent protein kinase which plays important roles in relaxation of vascular smooth muscle and inhibition of platelet aggregation (34). *FLT4* acts as a cell-surface receptor for vascular endothelial growth factor C (VEGFC) and vascular endothelial growth factor D (VEGFD), and plays an essential role in the development of the vascular network (35). It has been shown that VEGF and its receptor-related molecules are expressed in the brain tissues and reported to play protective during CM (36).

### Gene-based rare variant association

Because rare variants are known to play role in the variation of most complex traits, we applied optimal unified sequence kernel association test (SKAT-O), which combines burden and variance-component analyses (37) to the raw genotype GWAS dataset of Gambia, Kenya and Malawi populations (**see Material and Methods**). The SKAT-O analysis identified a total of six and nine nominally significant genes in Gambia and Malawi populations, respectively. These include nine long intergenic non-protein coding RNAs (LincRNAs), *MIR4282, GLYR1, NDNF, EPB41L2, ATP8A1* and *WASF3* (**Supplementary Table 3**). However, none of these genes were significant after correction for multiple testing.

### Functional networks and subnetworks of severe malaria resistance candidate genes

To investigate the functional interaction between all the candidate malaria resistance candidate genes identified in this study, we implemented network analysis (**see Materials and Methods**). Our global network generated 351 functional interactions between 268 genes. Topology analysis identified *ABO, HBB, HBD, HBE1, ATP2B4* as highly influential connector hub genes influencing at least two subnetworks/communities while *TRIM21* and *OR5F2* constituted independent communities. *MED22* and *OR551B6* constituted provincial hub genes (**Fig 3**).

**Fig. 3.**
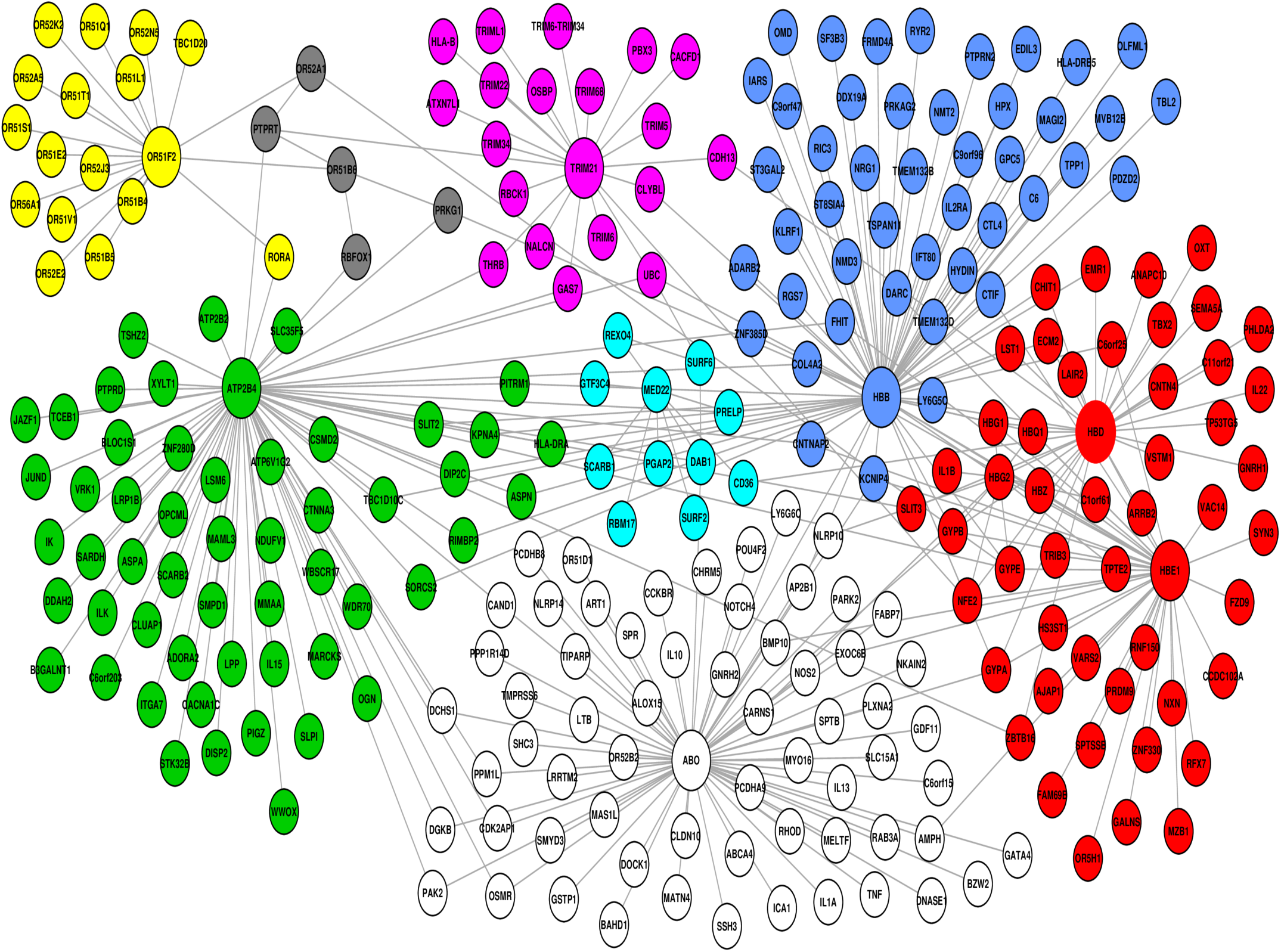
Network generated from predominant severe malaria protective candidate genes, comprising of 351 interactions between 268 nodes. Topology analysis identified *ABO, HBB, HBD, HBE1, ATP2B4* as highly influential connector hub genes influencing at least two subnetworks/communities while *TRIM21* and *OR5F2* constituted independent communities. *MED22* and *OR551B6* constituted provincial hub genes.

### Molecular functions and pathways of candidate severe malaria resistance genes

To test whether the genes predicted by the three functional mapping strategies overlapped in functional gene sets and pathways, we conducted gene enrichment analysis implemented in FUMA (20) using MsigDBc5 (38) gensets as background (**see Material and Methods**). The gene enrichment analysis identified several shared biological functions linked to erythrocyte-related pathways including three gene ontology (GO) cellular components, eight GO molecular functions and fourteen GO biological processes (**Table 2**). The implicated cellular components include haptoglobin-haemoglobin complex (p = 7.6e-8), haemoglobin-complex(p = 7.63e-8) and cytosolic-part (p = 6.73e-3). The enriched molecular functions include haptoglobin binding (p = 4.87e-8), oxygen carrier activity(3.8e-7), oxygen binding (p = 4.87e-5) and other activities related to haemoglobin functions. The shared biological activities include oxygen transport (p = 4.22e-6), gas transport (p = 4.87e-6), hydrogen peroxide catabolism (p = 9.08e-5), protein hetero-oligomerization (p = 2.86e-3), protein complex-oligomerization (p = 4.23e-3), interferon gamma mediated signalling pathways (p = 3.09e-2) and blood coagulation (p = 3.09e-2).

**Table 2.** Gene enrichment results of functionally annotated genes in malaria genomic risk loci identified by FUMA method.

| GO terms | GeneSet | N. genes | N. enriched genes | P-value | adjusted P-value | Genes |
| --- | --- | --- | --- | --- | --- | --- |
| Cellular components | <u>Haptoglobin-haemoglobin complex</u> | 11 | 5 | 8.91e-11 | 7.63e-8 | *HBB, HBD, HBG1, HBG2, HBE1* |
|  | <u>Haemoglobin complex</u> | 12 | 5 | 1.52e-10 | 7.63e-8 | *HBB, HBD, HBG1, HBG2, HBE1* |
|  | <u>Cytosolic part</u> | 239 | 7 | 6.73e-6 | 2.25e-3 | *HBB, HBD, HBG1, HBG2, HBE1, DBN1, SURF6* |
| Biological functions | <u>Haptoglobin binding</u> | 10 | 5 | 4.87e-11 | 8.02e-8 | *HBB, HBD, HBG1, HBG2, HBE1* |
|  | <u>Oxygen carrier activity</u> | 14 | 5 | 3.84e-10 | 3.16e-7 | *HBB, HBD, HBG1, HBG2, HBE1* |
|  | <u>Oxygen binding</u> | 36 | 5 | 6.87e-8 | 3.77e-5 | *HBB, HBD, HBG1, HBG2, HBE1* |
|  | <u>Molecular carrier activity</u> | 41 | 5 | 1.35e-7 | 5.56e-5 | *HBB, HBD, HBG1, HBG2, HBE1* |
|  | <u>Oxidoreductase activity acting on peroxide as acceptor</u> | 56 | 5 | 6.66e-7 | 2.19e-4 | *HBB, HBD, HBG1, HBG2, HBE1* |
|  | <u>Haemoglobin binding</u> | 7 | 3 | 8.69e-7 | 2.38e-4 | *HBB, HBD, HBE1* |
|  | <u>Antioxidant activity</u> | 85 | 5 | 5.35e-6 | 1.26e-3 | *HBB, HBD, HBG1, HBG2, HBE1* |
|  | <u>Tetrapyrrole binding</u> | 136 | 5 | 5.23e-5 | 1.08e-2 | *HBB, HBD, HBG1, HBG2, HBE1* |
| Biological Processes | <u>Oxygen transport</u> | 15 | 5 | 5.74e-10 | 4.22e-6 | *HBB, HBD, HBG1, HBG2, HBE1* |
|  | <u>Gas transport</u> | 19 | 5 | 2.20e-9 | 8.10e-6 | *HBB, HBD, HBG1, HBG2, HBE1* |
|  | <u>Hydrogen peroxide catabolic process</u> | 32 | 5 | 3.71e-8 | 9.08e-5 | *HBB, HBD, HBG1, HBG2, HBE1* |
|  | <u>Antibiotic catabolic process</u> | 50 | 5 | 3.74e-7 | 6.88e-4 | *HBB, HBD, HBG1, HBG2, HBE1* |
|  | <u>Drug catabolic process</u> | 108 | 6 | 8.05e-7 | 1.18e-3 | *CHIT1, HBB, HBD, HBG1, HBG2, HBE1* |
|  | <u>Cofactor catabolic process</u> | 66 | 5 | 1.52e-6 | 1.87e-3 | *HBB, HBD, HBG1, HBG2, HBE1* |
|  | <u>Protein hetero-oligomerization</u> | 133 | 6 | 2.73e-6 | 2.86e-3 | *HBB, HBD, HBG1, HBG2, HBE1, HIST1H3H* |
|  | <u>Protein complex oligomerization</u> | 551 | 10 | 4.60e-6 | 4.23e-3 | *TRIM21, HBB, HBD, HBG1, HBG2, HBE1, TRIM6, TRIM34, TRIM22, HIST1H3H* |
|  | <u>Antibiotic metabolic process</u> | 91 | 5 | 7.49e-6 | 6.11e-3 | *HBB, HBD, HBG1, HBG2, HBE1* |
|  | <u>Cellular detoxification</u> | 107 | 5 | 1.65e-5 | 1.21e-2 | *HBB, HBD, HBG1, HBG2, HBE1* |
|  | <u>Protein trimerization</u> | 54 | 4 | 2.00e-5 | 1.34e-2 | *TRIM21, TRIM6, TRIM34, TRIM22* |
|  | <u>Detoxification</u> | 122 | 5 | 3.11e-5 | 1.91e-2 | *HBB, HBD, HBG1, HBG2, HBE1* |
|  | <u>Interferon gamma mediated signalling pathway</u> | 70 | 4 | 5.60e-5 | 3.09e-2 | *TRIM21, TRIM68, TRIM34, TRIM22* |
|  | <u>Coagulation</u> | 335 | 7 | 5.89e-5 | 3.09e-2 | *HBB, HBD, HBG1, HBG2, HBE1, HIST1H3H, ADAMTS13* |
|  | <u>Oxygen transport</u> | 15 | 5 | 5.74e-10 | 4.22e-6 | *HBB, HBD, HBG1, HBG2, HBE1* |

In addition to genes within severe malaria genomic risk loci, we performed functional analysis and pathway analysis for the genes identified by the gene-based GWAS using Database for Annotation, Visualization and Integrated Discovery (DAVID) method (39) and Pascal (22), respectively(see Material and Methods). The DAVID analysis yielded eight functional categories, the majority of which are linked to malaria pathogenesis (**Table 3**) including GPCR signalling, membrane/transmembrane proteins, Na+/K+ transporting ATPases, cell adhesions, haemoglobin related functions, calcium signalling and actin binding activities.

**Table 3:**
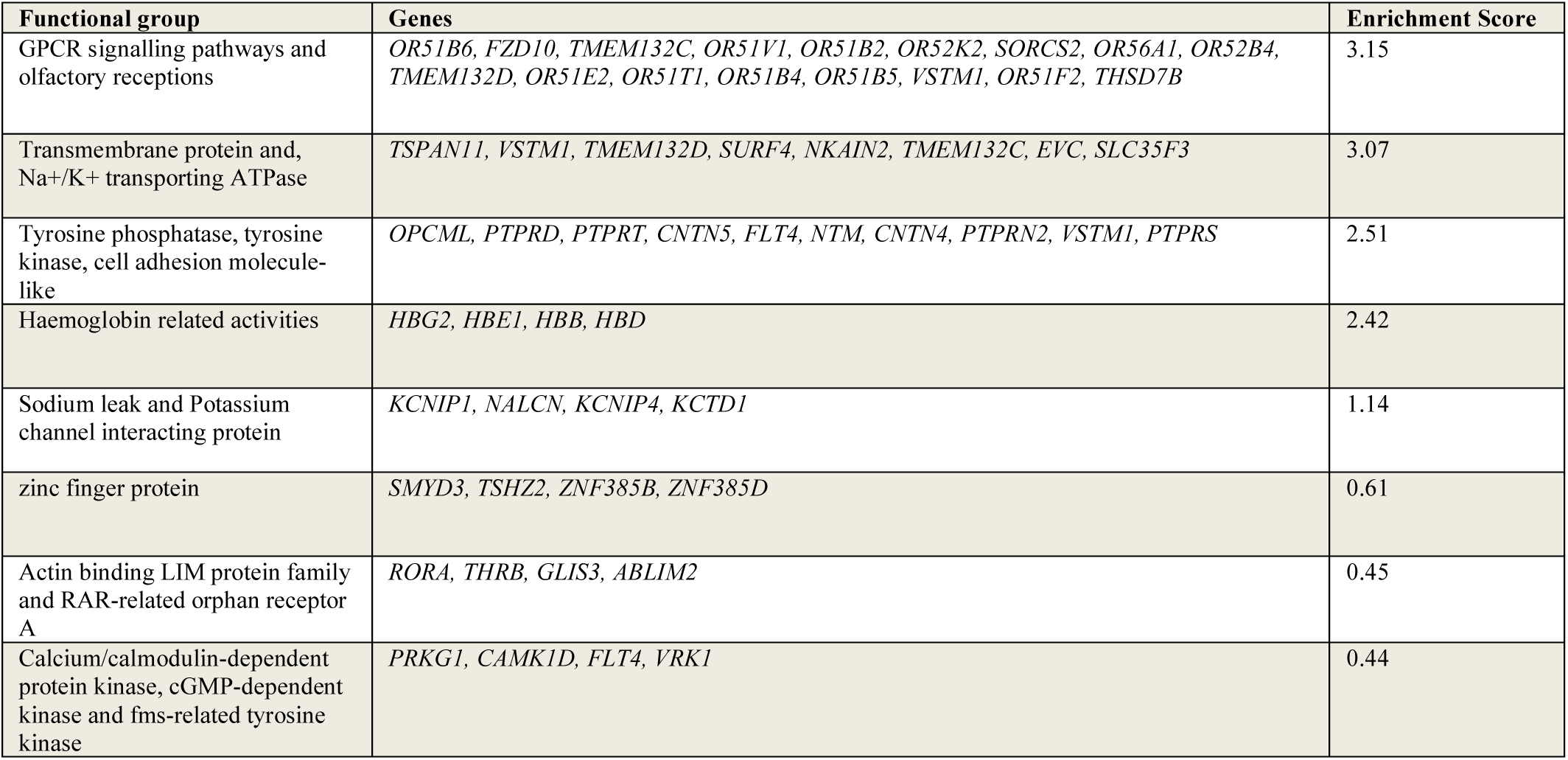
Functional categories of genes identified by gene-based GWAS analysis grouped using DAVID method.

Similarly, Pascal analysis implicated eleven significant pathways the majority of which are linked to malaria pathogenesis in RBCs, vasculatures and brain (**Fig 4A**). These include G protein-coupled receptor signalling (p = 7.88e-15), haemostasis (p = 4.52e-10), neuronal system (p = 1.25 e-9), axon guidance (p = 5.93e-8), calcium signalling (p = 1.10e-7), chemical transmission across synapses (p = 1.75 e-7), immune system (p = 2.61e-6), signalling by Rho GTPase (p = 1.45 e-5) and tight junction (p = 3.57 e-5). Furthermore, differential expression of genes implicated by Pascal method showed significant enrichment in blood vessels **(Fig 4B)**.

**Fig. 4.**
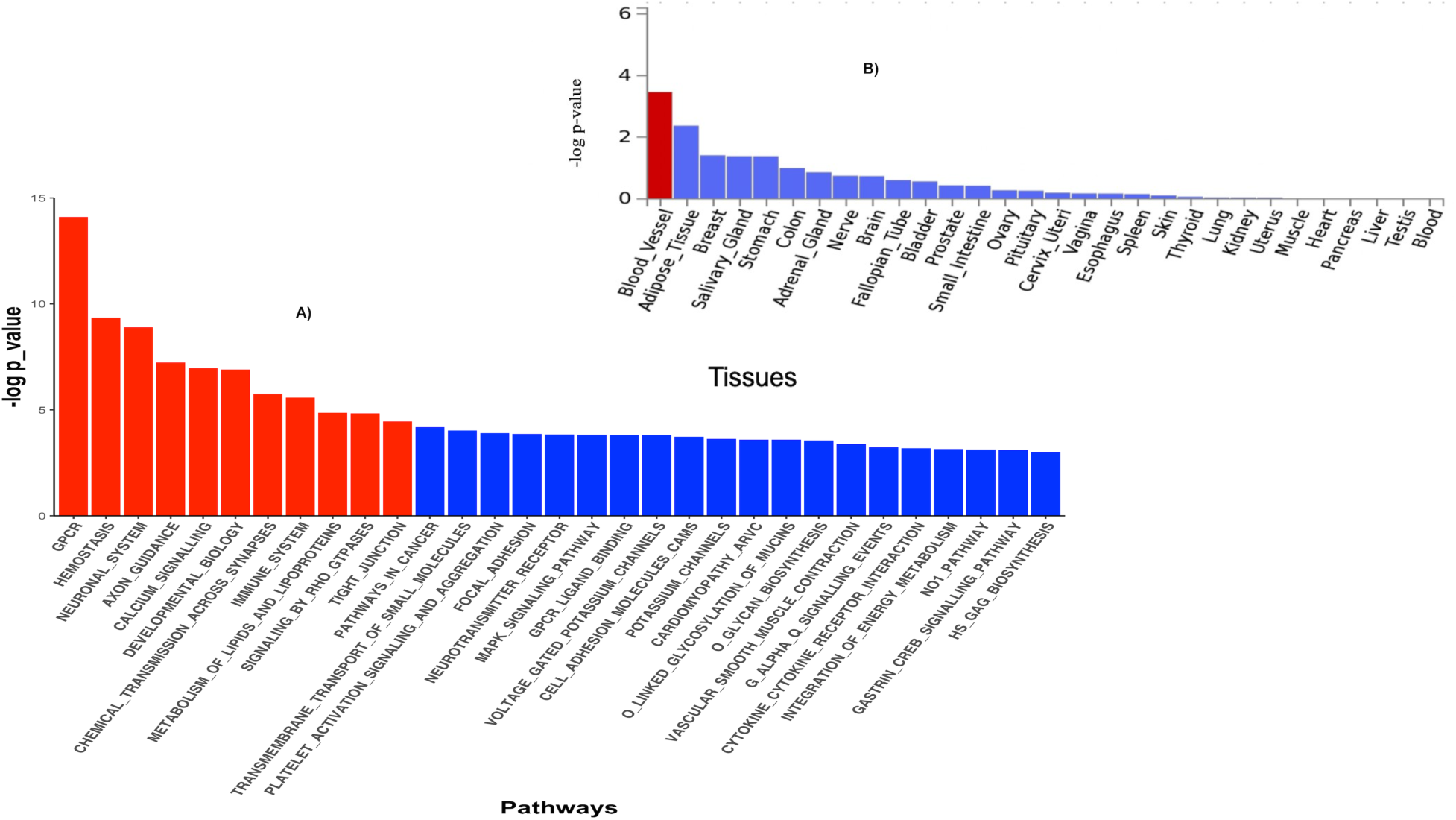
**A)** Pathway scores obtained from pascal analysis. Significant Pathways at Bonferroni corrected P-value ≤ 0.05 are coloured in red. **B)** Tissue specific gene expression and shared biological functions of genes identified by pascal in GTEx v8 30 general tissue types. Input genes were tested against each of the precomputed differentially expressed sets using the hypergeometric test. Significant enrichment at Bonferroni corrected P-value ≤ 0.05 are coloured in red.

### Population genetic structure of malaria resistance candidate genes

We noted that the minor allele frequencies (MAF) of the SNPs residing in the identified genes are generally higher in the three malaria populations compared to 20 ethnic groups (**Supplementary Fig 1 and 2**). We further observed that the proportion of pathogenic SNPs in a total of eighteen genes is much higher in the three malaria endemic populations compared to other populations (**Supplementary Data 6 and 7**). These include *TRIM* family genes such as *TRIM21, TRIM22, TRIM68, TRIM6-TRIM34 and TRIM34* in which the pathogenic SNP proportion ranges from 13.3–25%; olfactory receptors genes such as: *OR51B4, OR51B6, OR51B2, OR56A1, OR51L1, OR52K2 and OR51E2* in which the pathogenic SNP proportion ranges from (27.3 –100%).

The principal component analysis based on the SNPs residing in the identified genes effectively clustered all the populations according their ancestry (**Fig 5**).

**Fig.5.**
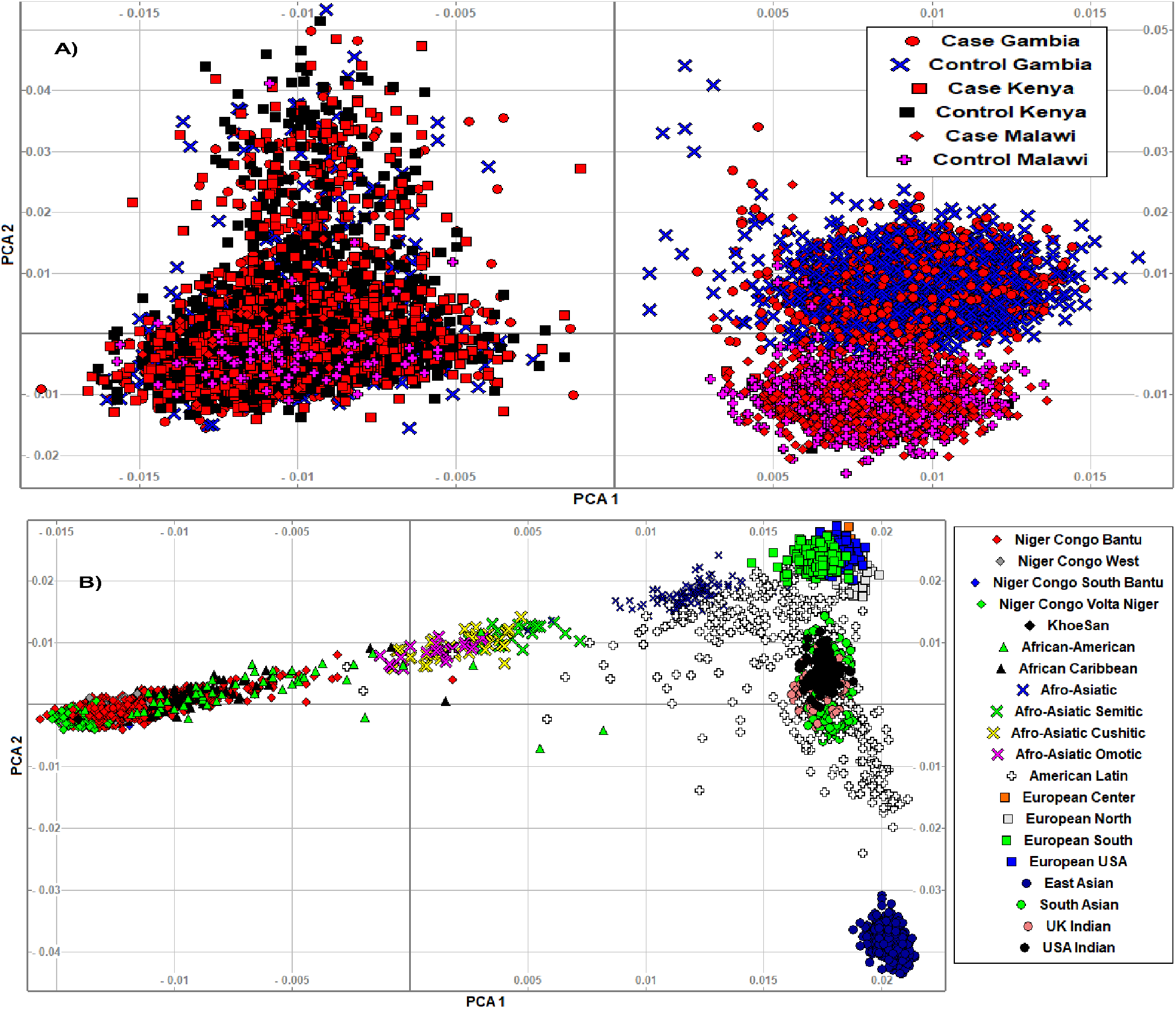
**A)** We clustered the merged malaria GWAS dataset containing only SNPs residing in the identified malaria resistance candidate genes (N = 10578 samples, 15,675 SNPs) using smartpca software. The three populations and their case/control status were indicated by different colours and symbols. **B)**. We clustered the AGVP dataset containing only SNPs residing in the identified malaria resistance candidate genes (N = 4932 samples, 93,5549 SNPs) in to sub-regions/populations using smartpca software. The populations were indicated by different colours and symbols.

## Discussions

In this study, we applied statistical functional analytic method to the largest ever severe malaria susceptibility GWAS dataset and identified the well-known malaria resistance loci and a number of novel genes that can guide future functional experiments. We noted that severe malaria resistance is attributed to multiple genes and pathways linked to malaria pathogenesis during blood stage life cycle of the parasite including merozoite invasion, parasite growth, cytoadherence, and signal transduction. The genes that were identified by our three mapping strategies might have equal importance; genes that were identified by positional mapping may act at protein level through structural changes while the genes identified by eQTL and chromatin interactions exert their influences through quantitative changes at gene expression levels (20).

The fact that the functionally mapped genes are clustered on chromosome 11p15 is consistent with our recent work in which we reported the disproportionate concentration of SNP-heritability on chromosome eleven. This might reinforce the need for targeting this chromosome in the future severe malaria susceptibility studies (11). We noted that in addition to the sickle trait gene *(HBB)*, our mapping strategies identified other members of beta globin gene cluster that cause various forms of beta-thalassemia (*HBE1, HBD, HBG* and *HBG2*).

Our network analysis showed that all these genes constituted hub with which several other genes are connected; which reaffirm the importance of homeopathies in resistance against severe malaria. Beta-thalassemia and other hemoglobinopathies are thought to confer protection against severe malaria by suppressing the parasite growth and by mitigating associated pathogenic effects (40). The proposed protective mechanisms of hemoglobinopathies against severe malaria has been extensively reviewed elsewhere (40, 41). In addition to the beta globin gene cluster, the Tripartite motif (*TRM)* containing gene family (*TRIM68* and *TRIM21*) identified in this locus are known to play critical role in down regulating Toll-like receptors(TLR)- and Rig-like receptors (RLR)-induced responses and protect from autoimmune and inflammations (42, 43). Mal-adapted inflammatory reactions is one of the hall mark pathogenic pathways in severe malaria (44, 45). The olfactory receptors super-family genes (*CCKR, OR51F2* and *OR51L*) identified in this locus might involve in G protein-coupled receptors(GPCR) signalling activities which is important in blood stage life cycle of *P. falciparum* (46). However, it is also possible that these genes were detected because of their abundance and close proximity to with globin gene cluster(47). We also be noted that the genes in the *TRM* family and olfactory receptors super-family contain higher proportion of pathogenic SNPs in the three-malaria endemic population compared to the global populations. The majority of the well-known malaria protective genes have deleterious variants and were evolved under balancing selections [42].

Our eQTL mapping identified *ADAMTS13* gene on chr9q34 outside the malaria genomic risk locus which would not have been identified by the conventional SNP mapping approach (20). *ADAMTS13* is a zinc-containing metalloprotease enzyme that cleaves, von Willebrand factor vWF, a large protein derived from endothelial surface and megakaryocytes which plays a crucial role in basic haemostasis (48, 49). Following activation of endothelial cells, vWF is directly released in to plasma and basement membrane or is stored in Weibel–Palade bodies (WPBs) from where it is released by regulated secretion to promote adhesion of platelets at the sites of vascular injury and facilitate vascular healing (49). However, abnormal accumulations of vWF caused by deficiency of plasma *ADAMTS13* trigger intravascular platelet aggregation and micro thrombosis leading to a vascular disease, Thrombotic Thrombocytopenic Purpura (TTP) (48). Indeed, recent works have linked the platelet-mediated clumping of infected erythrocytes in microvasculature during cerebral malaria with increased level of VWF in plasma caused by mutations in *ADAMTS13* genes (50–52).

In the same genomic locus, our eQTL functional mapping identified surfeit gene cluster, metalloprotease genes linked to epithelial adhesions and blood coagulations. *SURF4* gene has been implicated in epithelial cell adhesion trait (53), *MED22* gene is linked to VWF factor/factor VIII level measurement (54) and *SURF6* has been implicated in epithelial ovarian cancer (55). Our network analysis showed that *MED22* forms a central hub to which the rest of surfeit gene cluster are connected. This may suggest that the greater importance of *MED22* gene compared with the other members of the cluster. However, further comprehensive studies are needed to better understand the association of these genes with SM resistance. In the remaining genomic risk loci, the well-known genes including *ATP2B4* (<u>chr1q32</u>), *FREM3, GYPE* and *GYPB* (chr4p31) were replicated and few additional genes were identified. The glycophorin gene cluster, *GYPA* and *GYPB* encode the MNS blood group system and are host-erythrocyte receptors for *P. falciparum*; suggesting that polymorphisms in these genes play protective role by interfering with the invasion processes [55]. *ATP2B4* variants may impair the parasite lifecycle in erythrocyte by affecting intracellular Calcium homeostasis (18, 56).

Novel genes identified in the loci include *BTG2* on chr1q32 and *B3GALNT1* on chr3q26. *BTG2* is a tumour suppressor gene known to be linked to RBC related traits including MCHC level, RBC distribution and reticulocyte count (57). *B3GALNT1* encodes globoside blood group system which is determined by P antigen (58). Globoside/P antigen is the most abundant neutral glycolipid in the erythrocyte membrane and has been recognized as a cellular receptor for parvo-B19 virus (59). Individuals lacking this receptor are resistant to parvo-B19 virus and uro-pathogenic *E.coli* infections (59, 60). Further investigations are needed to establish the link between these genes and SM resistance.

We noted that the mapped genes share cellular components including haptoglobin binding, haemoglobin complex and cytosolic part and several overlapped molecular functions and biological processes linked with the blood stage life cycle of the parasite. *P. falciparum* spends most of its lifecycle within RBCs, where it undergoes multiple rounds of invasion, growth, replication and egress; causing the signs and symptoms of malaria (44, 61). The majority of the classical haemoglobin variants confer protection against severe malaria by restricting invasion process and intraerythrocytic growth of the parasite (40). Haptoglobin is an acute phase glycoprotein present in human plasma. It forms stable complexes with extracellular haemoglobin that is released from lysed RBCs and thereby curtail the haemoglobin-induced oxidative tissue damage (62).

*P. falciparum* ingest the host cell cytosol to obtain nutrients and space for growth in the RBCs (63). A recent study showed that the host cytosol uptake process is mediated by parasite’s protein called *VPS45* (64). The fact that the identified candidate genes in this study were enriched in the cytosol part of the cellular component might suggest that these genes might arrest the nutrient uptake of the parasites and thereby confer protection against their pathogenic effects. In addition to haemoglobin related functions, some of the candidate genes were enriched in other pathways linked to malaria pathogenesis including blood coagulation related processes malaria (65, 66) and interferon gamma mediated signalling pathways (67, 68). This may suggest that the host genetic factors might interfere with parasite development and its pathogenic outcomes at multiple levels to confer protection the life-threatening form of malaria.

Our gene-based GWAS analysis replicated the well-known malaria resistance candidate genes in the genomic risk loci and identified several genes across the genome. Genes with top scores encode for different malaria relevant functions such as: regulation of inflammation (*CSMD1*), neural adhesion *(CNTN4, PCSK5, CDH13, TMEM132*), vascular epithelial development (*FLT4*) and protein kinases (*PTPRT, PRKG1*). Furthermore, functional analysis of the candidate genes yielded functional categories linked with the blood stage lifecycle of the parasite and associated pathologies. The top enriched functions include GPCR signalling, membrane/transmembrane proteins, [Na+/K+] transporting ATPases, Sodium leak and Potassium channel interacting proteins, cell adhesions molecules and Calcium/Calmodulin-dependent protein kinases (CDPKs). CDPKs have crucial functions in calcium signalling at various stages of the parasite’s life cycle and is proposed to be one of the potential drug targets against malaria (69). It has been shown that *P. falciparum* infection activates host signalling pathway involving protein kinase C (PKC) (70). Similarly, host GPCR signalling pathways have been shown to play vital roles in invasion, intraerythrocyte parasite development and egress processes (71, 72); suggesting the existence of substantial interactions between host membrane/transmembrane signalling and parasite signalling elements which might mediate the disease severity. Further studies are needed to decouple the host-parasite interface of signal transduction and explore the potential target for new therapeutics. Sodium leak and Potassium channel interacting proteins, [Na^+^/K^+^] transporting ATPases play critical role in maintaining electrochemical equilibrium in normal erythrocytes. However, upon invasion by trophoblast stage of the parasite, the ion pump-leak balance is perturbed; with increased leak rate and decreased pump rate resulting in a remarkable increase in [Na+] and decrease in [K+] in the erythrocyte cytosol (73, 74). This results in formation of a new permeability pathway (NPP) in the erythrocyte membrane which allow the transport of nutrients and waste products necessary for the parasite. The composition and physiological role of NPP has been reviewed elsewhere (75, 76). Studies have shown that both parasites driven proteins encoded by *clag3.1* and *clag3.2* (77, 78) and host benzodiazepine receptor mediate the formation of NPP (79, 80). Our result may suggest the existence of multiple host genes that are involved in mediating this process. Given that TPP can be a potential target for new therapeutics, further studies are needed to investigate the role of host and parasites genetics in mediating the channel and its pathophysiology.

In addition to the well-known pathways such as: haemostasis and immune system that play role in malaria resistance (13), our analysis identified novel pathways including cell adhesion molecules, Rho GTPase activities, tight junction, neuronal system and axon guidance. One of the key virulence factors of *P. falciparum* is its capacity to modify iRBCs to adhere to the endothelium of the vasculature and, thereby, sequester in capillaries and postcapillary venules in vital organs leading to severe disease manifestations (61). Adhesion phenotype is mediated by expression of *P. falciparum* erythrocyte membrane protein 1 (*PfEMP1*) on the iRBCs (81, 82). The binding of iRBCs with endothelium involve various adhesion molecules including CD36, ICAM-1, E-selectin and chondroitin sulfate A (CSA) that are variably expressed in different organs (83–85). The neural adhesion molecules identified in the current study might involve in receptor activities and their polymorphisms might play protective roles against SM. Furthermore, adhesion events have been shown to activate Rho kinase signalling pathway which is strongly implicated in various vascular diseases (86). The genes that are enriched in these pathways might provide protection against severe malaria by weakening the cytoadherence interactions and associated pathologies. Other genes that are enriched in neuronal system, axon guidance and tight junction might be linked with intra cerebral pathogenesis of SM (87). Furthermore, the candidate malaria resistance genes identified by gene-based GWAS were differential expressed in blood vessels; suggesting that the majority of the identified genes are likely counteract *P. falciparum* induced endothelial disfunctions in microvasculature and capillaries (44).

In conclusion, our functional mapping analysis identified 57 genes located in the known malaria genomic loci while our gene-based GWAS analysis identified additional 125 genes across the genome which can potentially guide future experimental studies. The identified genes were significantly enriched in malaria pathogenic pathways including multiple overlapping pathways in erythrocyte-related functions, blood coagulations, ion channels, adhesion molecules, membrane signaling elements and neuronal systems. Overall, our results suggest that severe malaria resistance trait is attributed to multiple genes that are enriched in overlapping pathways linked to severe malaria pathogenesis; highlighting, the possibility of harnessing new malaria therapeutics that can simultaneously target multiple malaria protective host molecular pathways. Further experimental studies are needed to validate the findings in the current study.

## Materials and Methods

### Description of the study datasets

We accessed a previous severe malaria GWAS datasets (16) (N = ∼11,000) of three African populations including Kenya, Gambia and Malawi from European Phenome Genome Archive (EGA) following the standard data access protocols outlined in (88, 89). Children with severe malaria cases were recruited on admission to hospital using definitions as per WHO guidelines for cerebral malaria (Blantyre coma score < 3 in children or Glasgow coma score < 11 in adults), severe malarial anaemia (haemoglobin < 5 g/100 mL or haematocrit < 15%) and other malaria-related symptoms (90). Control samples were obtained from representative of the ethnic groups of the cases or in some study sites from the local population (89). The samples were genotyped on Illumina Omni 2.5Marray and QC filtered as described in (14). In addition to the genotype dataset, we obtained a set of severe malaria susceptibility GWAS summary statistics (N = 17,000) meta-analysed across eleven population in Africa, Oceania and Asia from (18). The dataset contained information on GWASs of individual study populations and their meta-analysis. We additionally accessed a merged data set from 1000 Genomes Project and African Genome Variation Project(AGVP)(91) of which 20 world-wide ethnic groups (**Supplementary Table 4**) have been grouped following ethno-linguistic information (92).

### Functional mapping and annotations

We used the meta-analyzed malaria GWAS summary statistics (N = 17,000 samples, 17million SNPs) across eleven populations(18) for functional mapping and annotations. We implemented FUMA(20), a pipeline that determines genomic risk loci and prioritize potential causal genes by incorporating information from multiple sources including GTEx (93), ENCODE (25), Roadmap Epigenomics Project (26) and chromatin interaction information (27). Briefly, a pre-calculated LD structure based on 1000 Genome version 3 of the African population was used to determine the risk loci and independent significant SNPS from the GWAS summary statistics data. Independent significant SNP was defined as a genome-wide significant SNP (P-value < 5e-8) within LD threshold of r^2^ > 0.6. Other nominally significant SNPs (p< 0.05) that are in LD (r^2^ < 0.6) with independent significant SNPs were designated as candidate SNPs and included for annotation analysis. From identified independent significant SNPs, lead SNPs were selected at LD threshold of r^2^ > 0.1. SNPs in close proximity (< 250 k) were considered as a single locus and thus, each genomic locus can contain multiple independent significant SNPs and lead SNPs.

We then implemented three gene mapping strategies including positional mapping, expression Quantitative Trait Locus (eQTL) mapping, and chromatin interaction implemented in FUMA(20). Positional mapping was performed by ANNOVAR tool (94) using Ensembl (build 85; http://www.ensembl.org/) dataset. A maximum distance of 10kb window size upstream and downstream was used to map SNPs to genes. SNPs filtering was carried out based on CADD score(95), RegulomeDB score (96) and 15-core chromatin state (97).

eQTL mapping was performed for genes within 1 Mb of the most significant variant using datasets that contain eQTL information related to severe malaria such as brain and blood. These include:PsychENCODE(98), GTExv8 (93), BRAINEAC (99), DICE (100), eQTLGen (101) (and Blood eQTLbrowser (102) and scRNA_eQTLs(103). Chromatin interaction mapping was performed using dataset including Hi-C data of 21 tissues/cell types obtained from GSE87112. (104), Hi-C loops from Giusti-Rodriguez et al. 2019 (105) which contains pre-processed enhancer-promoter and promoter-promoter interactions based on Hi-C data for adult and foetal human brain samples(105), Hi-C based data from PsychENCODE (98) which is composed of Enhancer-Promoter links based on Hi-C and Promoter anchored Hi-C loops and Enhancer-Promoter correlations from FANTOM5 (106). We restricted our analysis to tissues related to severe malaria pathogenesis. Genes prioritized by any of these three strategies were considered as potential causal genes. To gain insights in to the biological functions of prioritized genes, we performed gene enrichment analysis using an hypergeometric test in which gene-sets obtained from MsigDB (38) and WikiPathways (107) were used as background genes. We further tested differential gene expression values on 54 tissues obtained from the GTEx (93) as indicted FUMA (20).

### Gene-based genome-wide association analysis

Considering the polygenic nature of severe malaria susceptibility trait (11), we applied Pascal (22), a gene-based GWAS analysis that can aggregate all SNPs within a gene and thus, capture modest effects. Briefly, Pascal sum of chi-squared statistics (SOCS) analysis was applied to nominally significant GWAS SNPs (p-value < 0.05) and to compute the corresponding gene scores (p-values) in 50kb upstream and downstream window. LD information for estimation of correlation structure was obtained from African dataset in 1000G phase 3 (108). We further categorized the prioritized genes in to different functional groups using DAVID tools (39). Significant genes were subjected to differential gene expression analysis implemented in FUMA software using 54 tissues obtained from the GTEx (93). Pathway scores were computed by combining the scores of genes that belong to the same gene-set. The analysis doesn’t require a gene score threshold and incorporates weakly associated genes for pathway enrichment. Gene-fusion parameter was set to 1Mb so that all pathway-member genes within 1Mb apart were fused together. Genes in HLA-region and those containing more than 3000 SNPs were removed as outlined in Pascal documentations (22).

### Gene burden and rare-variants association tests

Given that the GWAS assumption is based on common variant common disease hypothesis, GWAS approach always miss potential association signal from rare variants. To examine the contribution of rare variants, we applied optimal unified sequence kernel association test (SKATO), which combines burden and variance-component analyses (37) to the GWAS dataset of Gambia, Kenya and Malawi populations. Briefly, we aligned the VCF files including Gambia (N = 4920 samples, 1.6 million SNPs), Malawi (N = 2560 samples, 1.6 million SNPs) and Kenya (N = 3143 samples, 1.6 million SNPs) to GWAS dataset to 1000 Genome v-3 reference haplotypes using Genotype Harmonizer (109) and removed SNPs with position and strand mismatches and phased using SHAPEITv2 (110). We performed imputation using impute 2 (111) and obtained ∼ 20 million from each population. After removal SNPs with low genotype rate and imputation accuracy, we retained ∼15,000 SNPs in each population. We then applied SKAT-O test to the quality filtered data following the procedure outlined in SKAT package (37).

### Network analysis

We performed a network analysis to investigate the functional interaction between malaria resistance candidate genes. Briefly, we obtained functional interaction network from of the identified candidate malaria resistance genes using Multiple Association Network Integration Algorithm (geneMANIA) tool (112). Using this information, we computed network parameters including degree, betweenness and closeness centrality metrics to evaluate the topology of nodes(genes) and edges(interactions) in the network using networkX (113) and R igraph packages (114).

Closeness measures the average distance from the node to all other nodes in the network, indicating which nodes represent a greater “risk” (maximally close with lowest sum of edge weights) for eliciting other nodes. Betweenness measures the number of times that a node lies on the shortest path between two other nodes, indicating which nodes serve as a “hub” between other nodes (114). The degree of a node is described as the number of direct connections it has with other nodes within the network. Low degree nodes usually connect to nodes within their local community whereas high degree nodes usually extend to the neighbouring community (114). Using the centrality scores, we quantified node centrality to identify hub genes by investigating the contribution of the edges and the weight of the edges towards node centrality. The hubs genes make strong contributions to the subnetwork and/or global network integrity. Connector hubs and provincial hubs refers to nodes that link other nodes across different communities and local communities, respectively.

### Population genetic structure of malaria-specific resistance candidate genes

We performed the population genetic structure of the identified genes in malaria endemic populations (Kenya, Gambia and Malawi) and global populations of 20 ethnic groups obtained from African Genome Variation Project (AGVP) (91). We mapped a total of 14,106,476 SNPs to the identified candidate genes using dbSNP database. We merged the quality filtered GWAS datasets of the three malaria endemic populations using PLINK software (115) and retained dataset that contains the SNPs that are mapped to the identified genes (N = 10578 samples, 15,675 SNPs). We performed basic quality control, removed structural variants and ambiguous SNPs using PLINK software (115) as described elsewhere (116). We retained AGVP dataset containing only the SNPs that are mapped to the identified candidate genes (N = 4932 samples, 93,5549 SNPs). We then partitioned these datasets in to a total 23 different Ethnic groups (20 from AGVP and 3 from malaria endemic populations) based on population or country labels information. We clustered the merged malaria GWAS dataset and the AGVP dataset containing only SNPs residing in the identified genes in to sub-regions/populations using smartpca software (117). For each ethnic group, we computed various population genetics analysis including minor allele frequency (MAF) at various bins and gene-specific in SNPs MAF by aggregating MAF of all associated SNPs within gene from dbSNPs. We finally computed proportion of pathogenic SNPs within the candidate genes using ANNOVAR software (94) by computing the number of SNPs reported to be pathogenic over total numbers associated SNPs within gene from dbSNPs.

## Data Availability

The GWAS summary statistics used in this study can be found at https://www.malariagen.net/sppl25/

## Acknowledgements

This work was supported through the DELTAS Africa Initiative [grant 107740/Z/15/Z]. The DELTAS Africa Initiative is an independent funding scheme of the African Academy of Sciences (AAS)’s Alliance for Accelerating Excellence in Science in Africa (AESA) and supported by the New Partnership for Africa’s Development Planning and Coordinating Agency (NEPAD Agency) with funding from the Wellcome Trust [grant 107740/Z/15/Z] and the UK government. The authors would also like to thank the National Research Foundation of South Africa for funding (NRF) [grant # RA171111285157/119056] and The Centre for High-Performance Computing (CHPC, http://www.chpc.ac.za) for computing resources This study makes use of data generated by MalariaGEN. A full list of the investigators who contributed to the generation of the data is available from http://www.malariagen.net. The views expressed in this publication are those of the author(s) and not necessarily those of AAS, NEPAD Agency, Wellcome Trust or the UK government.

## Authors contributions

DD designed, performed the data analysis and drafted the manuscript, FA, PK, NK, WT, HG and LG contributed in data analysis and revision of the manuscript, EC supervised the work

## Supplementary Figures and Tables

**Supplementary Fig 1.**
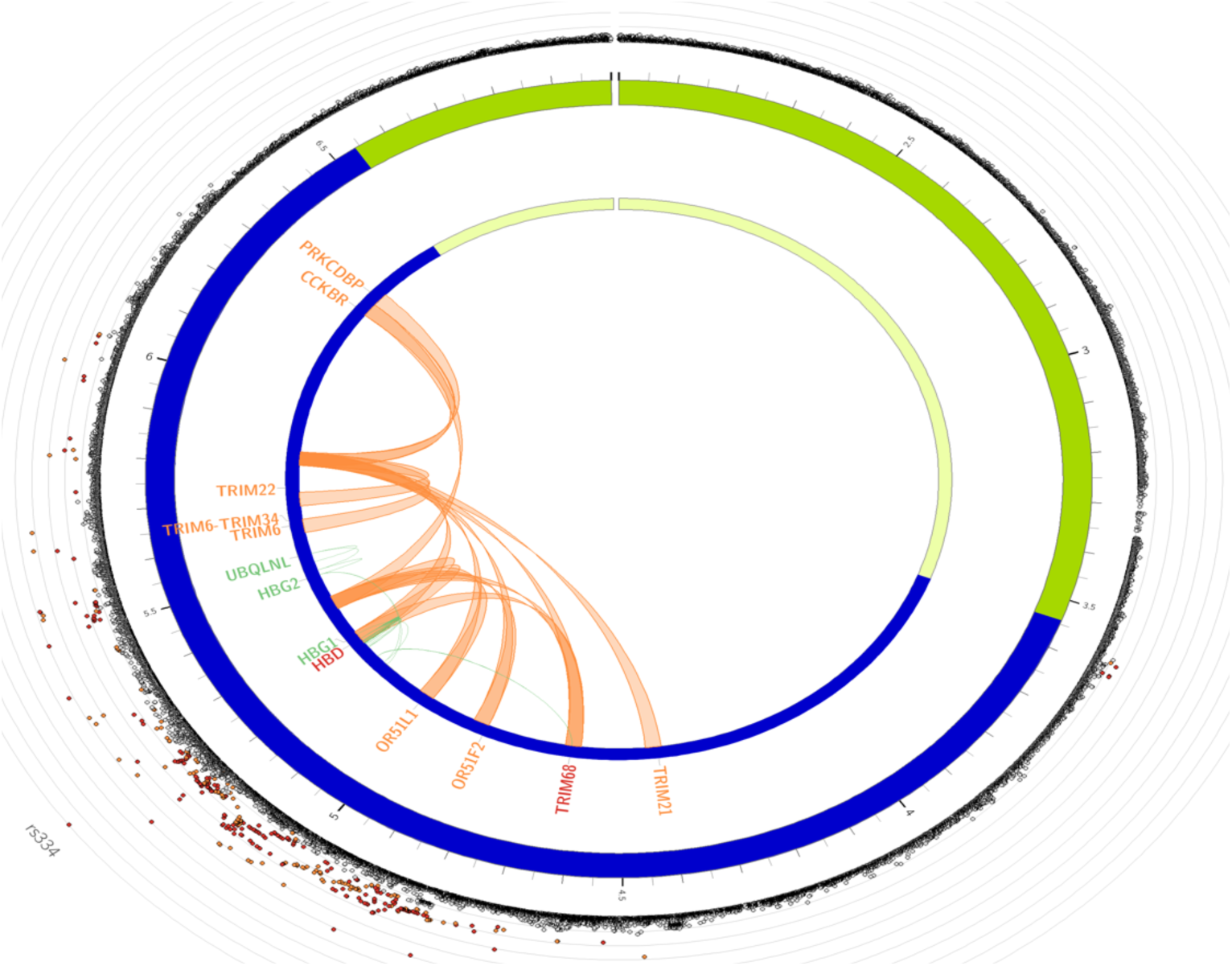
Chromatin interactions and eQTLs of severe malaria resistance candidate genes on chr 11 risk locus. The most outer layer is the Manhattan plot displaying SNPs with P-value < 0.05. Candidate SNPs are coloured based on the highest r^2^ to one of the independent significant loci (red: r^2^ > 0.8, orange: r^2^ > 0.6). Other SNPs are coloured in grey. The outer circle is the chromosome coordinate and genomic risk loci are highlighted in blue. Genes mapped by either Hi-C or eQTLs are shown on the inner circle. Genes identified by chromatin interaction and eQTLs are coloured orange and green respectively while genes mapped by both are coloured red.

**Supplementary Fig 2.**
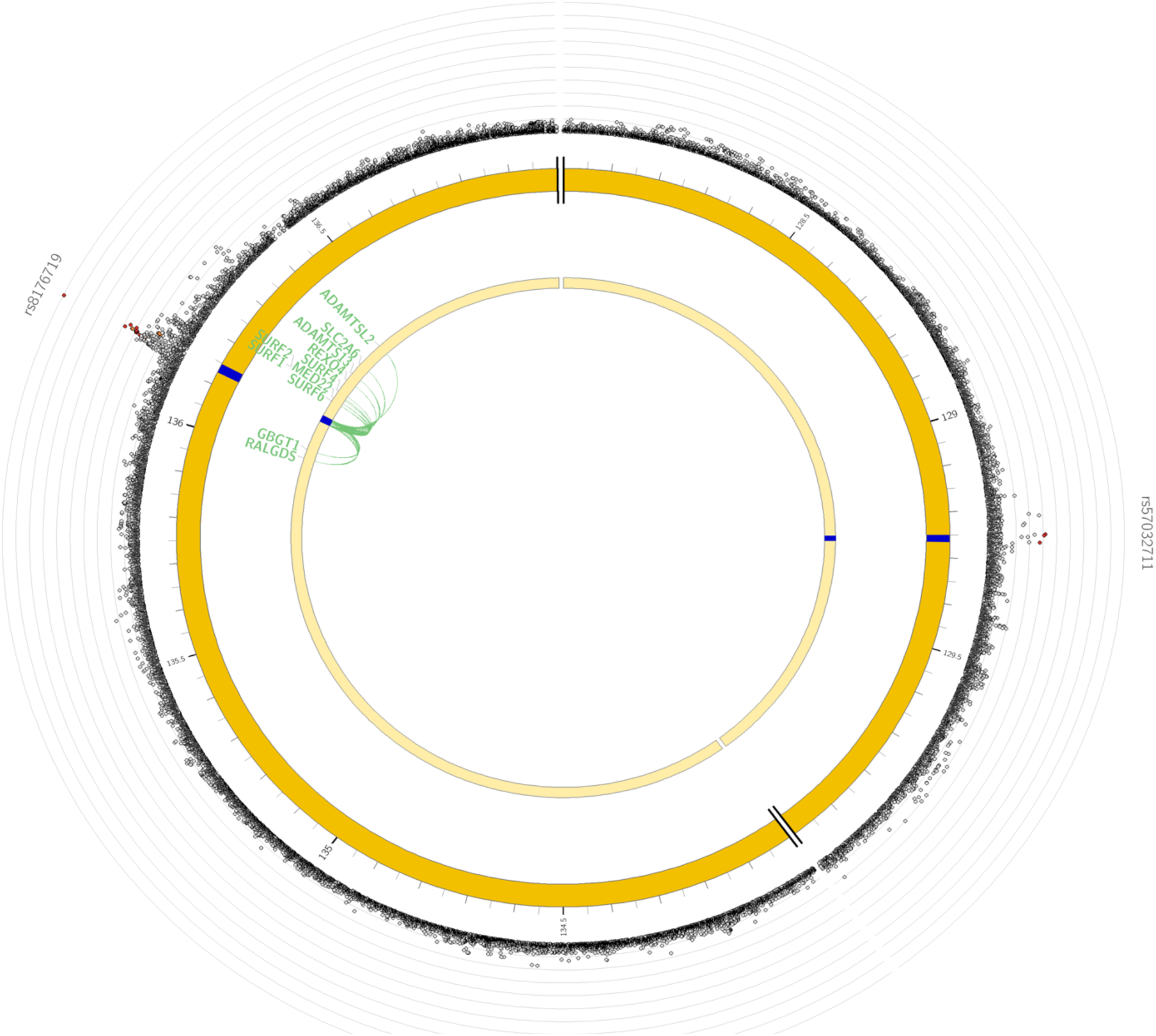
Chromatin interactions and eQTLs of severe malaria resistance candidate genes on chr 9 risk locus. The most outer layer is the Manhattan plot displaying SNPs with P-value < 0.05. Candidate SNPs are coloured based on the highest r^2^ to one of the independent significant loci (red: r^2^ > 0.8, orange: r^2^ > 0.6). Other SNPs are coloured in grey. The outer circle is the chromosome coordinate and genomic risk loci are highlighted in blue. Genes mapped by either Hi-C or eQTLs are shown on the inner circle. Genes identified by chromatin interaction and eQTLs are coloured orange and green respectively while genes mapped by both are coloured red

**Supplementary Fig 3.**
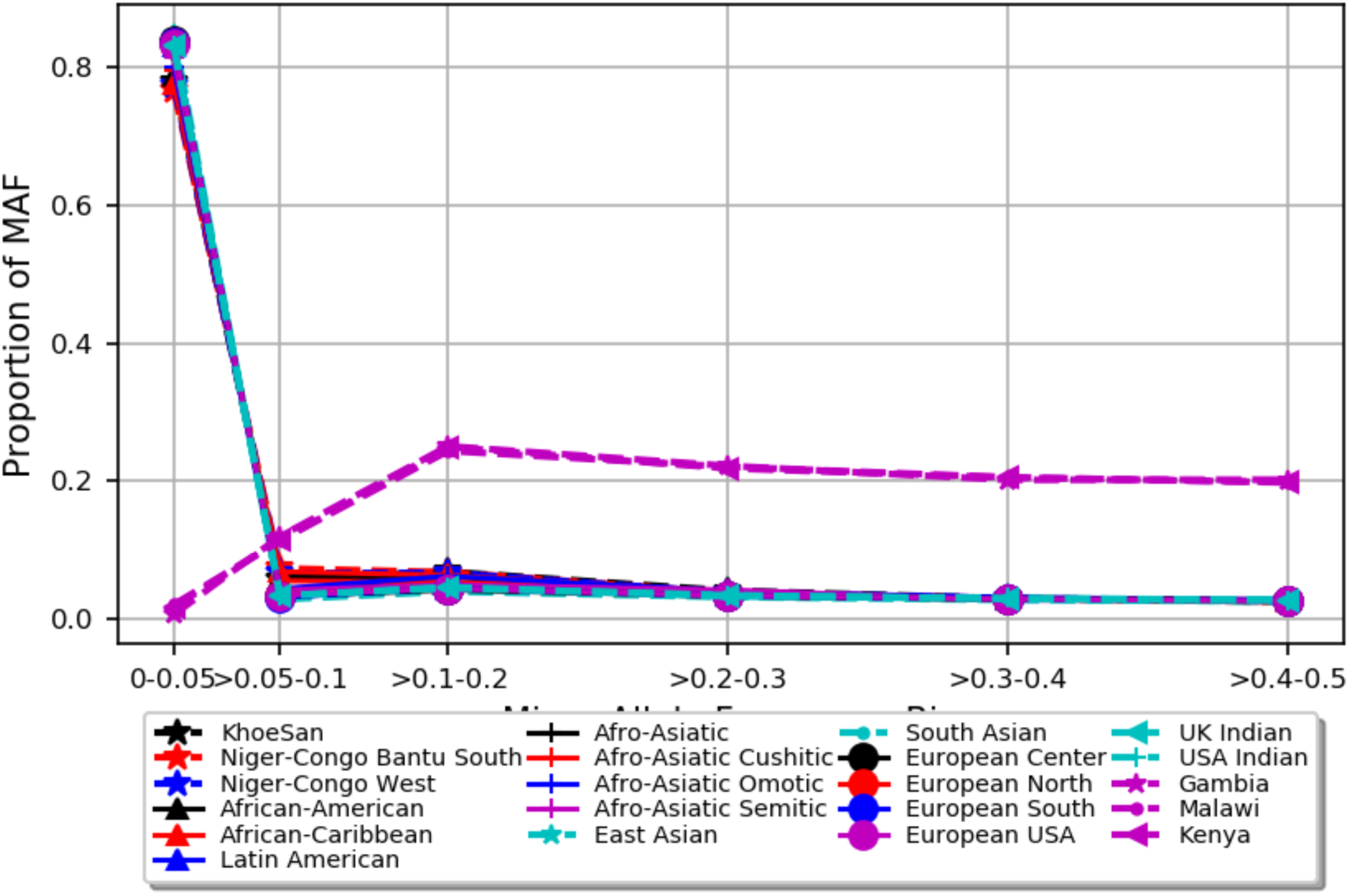
Minor allele frequency bins of SNPs mapped to severe malaria resistance candidate genes in three malaria endemic populations (Gambia, Malawi, Kenya) and global populations of 20 ethnic groups. Y-axis represent allele frequency, X-axis represent different MAF bins. Populations were represented by different colors and symbols

**Supplementary Fig 4.**
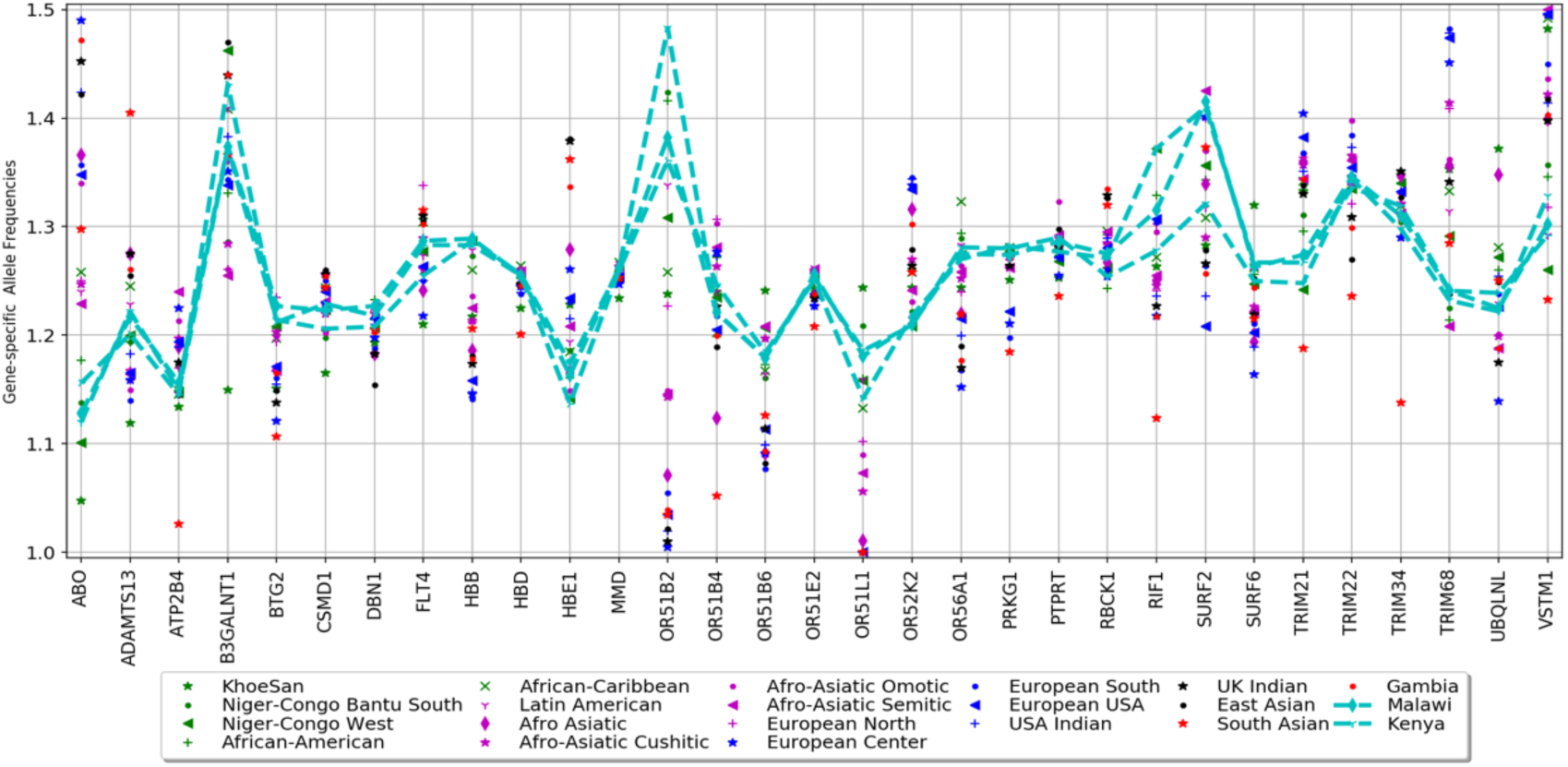
Gene specific MAF of SNPs mapped to severe malaria resistance candidate genes in three malaria endemic populations (Gambia, Malawi, Kenya) and global populations of 20 ethnic groups. Y-axis represent gene specific allele frequency, X-axis represent genes. Populations were represented by different colors and symbols

**Supplementary Table 1.** Positional enrichment of genes identified by FUMA method using MsigDB genes as background.

| Ensg | entrezID | HUGO | symbol | chromosome | Cytoband | Start | end | Biotype |
| --- | --- | --- | --- | --- | --- | --- | --- | --- |
| ENSG00000058668 | 493 | ATP2B4 | ATP2B4 | 1 | q32.1 | 203595689 | 203713209 | Protein coding |
| ENSG00000148296 | 6838 | SURF6 | SURF6 | 9 | q34.2 | 136197552 | 136203235 | Protein coding |
| ENSG00000148297 | 6837 | MED22 | MED22 | 9 | q34.2 | 136205160 | 136214986 | Protein coding |
| ENSG00000148290 | 6834 | SURF1 | SURF1 | 9 | q34.2 | 136218610 | 136223552 | Protein coding |
| ENSG00000148291 | 6835 | SURF2 | SURF2 | 9 | q34.2 | 136223428 | 136228045 | Protein coding |
| ENSG00000148248 | 6836 | SURF4 | SURF4 | 9 | q34.2 | 136228325 | 136242970 | Protein coding |
| ENSG00000132109 | 6737 | TRIM21 | TRIM21 | 11 | p15.4 | 4406127 | 4414926 | Protein coding |
| ENSG00000176925 | 119694 | OR51F2 | OR51F2 | 11 | p15.4 | 4842551 | 4843686 | Protein coding |
| ENSG00000244734 | 3043 | HBB | HBB | 11 | p15.4 | 5246694 | 5250625 | Protein coding |
| ENSG00000223609 | 3045 | HBD | HBD | 11 | p15.4 | 5253908 | 5256600 | Protein coding |
| ENSG00000196565 | 3048 | HBG2 | HBG2 | 11 | p15.4 | 5274420 | 5667019 | Protein coding |
| ENSG00000213931 | 3046 | HBE1 | HBE1 | 11 | p15.4 | 5289582 | 5526847 | Protein coding |
| ENSG00000184881 | 79345 | OR51B2 | OR51B2 | 11 | p15.4 | 5344541 | 5345582 | Protein coding |

**Supplementary Table 2.**
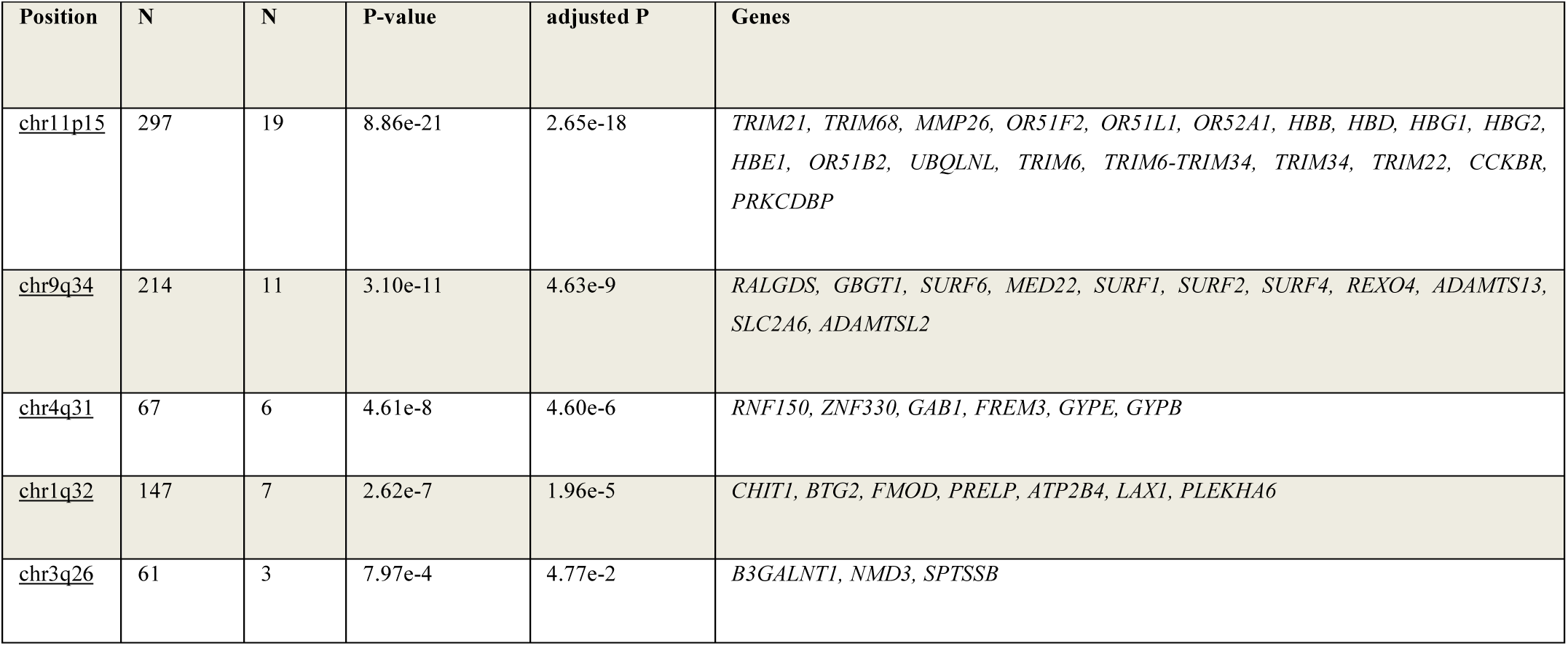
Candidate malaria resistance genes identified by both functional mapping and gene-based GWAS analysis

**Supplementary Table 3.** Nominally significant genes obtained from burden and rare variant analysis using SKAT package

| Populati<br>on | Gene | chro<br>moso<br>me | Cytoban<br>d | start | end | Biotype | P.value | N.Marker.Rare |
| --- | --- | --- | --- | --- | --- | --- | --- | --- |
| Gambia | *LOC105377246* | 4 | q13.1 | 58,976,071 | 58,984,194 | ncRNA | 1.72e-4 | 29 |
|  | *LOC100507464* | 8 | q11.21 | 49,496,763 | 49,512,234 | ncRNA | 9.68e-05 | 36 |
|  | *MIR4282* | 6 | q13 | 72967687 | 72967753 | lncRNA | 4.83e-4 | 7 |
|  | *GLYR1* | 16 | P13.3 | 4803203 | 4847288 | Protein<br>coding | 3.72e-4 | 139 |
|  | *LINC00520* | 14 | q22.3 | 55781132 | 55796731 | lncRNA | 9.70e-05 | 19 |
|  | *LOC105371381* | - | - | - | - | - | 1.84e-4 | 15 |
| Malawi | *ATP8A1* | 4 | P13 | 42408373 | 42657105 | Protein<br>coding | 4.91e-4 | 244 |
|  | *EPB41L2* | 6 | q23.2 | 130839347 | 131063322 | Protein<br>coding | 3.91e-4 | 207 |
|  | *LOC105376161* | 9 | q22.32 | 95,764,922 | 95,776,282 | ncRNA | 1.51e-4 | 9 |
|  | *LOC105377731* | 5 | q35.1 | 173,269,431 | 173,280,929 | ncRNA | 3.13e-4 | 29 |
|  | *NDNF* | 4 | q27 | 121035613 | 121073021 | Protein<br>coding | 9.53e-05 | 28 |
|  | *LINC00676* | 13 | q13.34 | 109,728,274 | 109,730,034 | lncRNA | 3.40e-4 | 10 |
|  | *LOC105370198* | 13 | q14.2 | 48,108,123 | 48,166,377 | ncRNA | 4.87e-4 | 122 |
|  | *LOC653786* | - | - | - | - |  | 4.53e-4 | 29 |
|  | *WASF3* | 13 | q12.13 | 26557683 | 26688948 | Protein<br>coding | 2.54e-4 | 193 |

